# Glucoregulatory and Inflammatory Trajectories in Pregnancy with Obesity: Secondary Analysis of a Randomized Exercise Trial

**DOI:** 10.64898/2026.10.21.26366153

**Authors:** Lisa T. Jansen, Simon Chung, Aaron R. Caldwell, Andrew W. Brown, Brittany N. Burgener, Audrey Martinez, Elisabet Børsheim, Aline Andres

## Abstract

**Objective:** To evaluate moderate-intensity exercise effects on glucoregulatory and inflammatory markers in pregnant women with obesity.

**Methods:** Secondary analysis of the Expecting Study (NCT02125149) included 263 sedentary pregnant women with pre-pregnancy BMI ≥30kg/m^2^ randomized to exercise (EX, n=157) or standard-of-care (SoC, n=106). Fasting biomarkers were assessed at 12.14±0.50, 24.19±0.68, and 36.03±0.58 weeks’ gestation. Linear mixed-effects models evaluated intervention and gestational effects at 24-36 weeks. Exploratory repeated-measures continuous glucose monitoring (CGM) included 18 participants (SoC=6, EX=12), evaluating 24-h/temporal glycemia and exploring inflammatory marker associations.

**Results:** Insulin, C-peptide, HOMA2-IR, *HOMA2-%B*, IL-6, and TNF-α increased with gestation, while *HOMA2-%S* and CRP decreased (all p<0.001). Exercise did not affect fasting glucose or inflammatory markers. CGM-wear yielded 12-15 days/trimester with >95% valid wear time; glycemia remained normoglycemic. By 36-weeks, SoC-metrics declined in 24-h and nocturnal glucose and time-in-range (TIR), with increased time-below-range (all p<0.022). TIR was inversely associated with IL-6 at 24-weeks (−0.23 [95%CI: −0.43,-0.02] p=0.035), and mean amplitude of glycemic excursion positively with TNF-α at 36-weeks (0.02 [95%CI: 0.00,0.04] p=0.033).

**Conclusion:** Exercise did not modify mid-to late-gestation fasting glucoregulatory or inflammatory profiles. High CGM-adherence and preliminary indications of distinct longitudinal glycemic patterns support further CGM evaluation in antepartum behavioral interventions.

**What is already known about this subject?:**

- Pregnancy and obesity increase insulin resistance and metabolic stress; exercise may improve maternal metabolic health.
- Continuous Glucose Monitoring (CGM) can detect glycemic patterns not captured by fasting measures but is understudied in pregnancies with obesity without diabetes.

**What are the new findings in your manuscript?:**

- Exercise did not alter fasting glucoregulatory or inflammatory trajectories despite substantial gestational metabolic adaptation.
- Exploratory CGM showed declining late-gestation glycemic metrics in the standard of care group, whereas these metrics remained stable in the exercise group.

**How might your results change the direction of research or the focus of clinical practice?:**

- Fasting biomarkers alone may not capture metabolic responses to antepartum exercise.
- Larger trials should evaluate CGM-derived glycemic patterns and clinically meaningful targets in pregnancies with obesity.

## Introduction

From 2016-2019, pre-pregnancy obesity increased by 11% and now affects 29% of women in the United States(1). Most U.S. Southern states exceed this national average, with Arkansas recording the nationwide steepest increase at 22%(1). This trajectory expands the number of women entering pregnancies with elevated metabolic load(2), as obesity amplifies the already substantial metabolic demands of pregnancy, and heightens the risk for adverse pregnancy outcomes(3).

Obesity and pregnancy independently challenge glucose regulation in distinct ways. While obesity drives chronic low-grade inflammation and persistent insulin resistance(4), pregnancy introduces a physiologic, temporary insulin resistance in mid-to late-gestation to prioritize glucose availability to the fetus(5). This adaptive insulin resistance is tightly regulated; however, when maternal obesity overlays this process, insulin resistance can be exacerbated from a physiologic to pathologic state, thereby increasing the risk for adverse maternal and infant health outcomes(6). Furthermore, hypertensive disorders in pregnancy are linked to excess maternal weight, increased levels of inflammatory markers, and glycemic variation(7). Lifestyle interventions, specifically exercise, harbor potential to ameliorate the metabolic strain placed on pregnant women with obesity(8, 9). Antepartum exercise interventions have demonstrated maternal metabolic health benefits, including limiting gestational weight gain across BMI categories(10), improving maternal insulin dynamics in women with healthy weight(11), managing glucose levels in gestational diabetes(12), and reducing the risk of gestational diabetes onset in women with overweight or obesity(13). Among women with excess weight, time spent in vigorous activity during early pregnancy has been linked to improved insulin sensitivity in late gestation(14). Nonetheless, fewer than 29% of pregnant women in the U.S. meet the physical activity guidelines at any gestational age(15). Understanding how new adoption of exercise influences glucose regulation in pregnancy with obesity is essential to inform future interventions aimed at preventing adverse health outcomes in both mothers and infants. Therefore, we tested a *de novo* moderate-intensity, mixed-modality exercise intervention in women with obesity and sedentary behaviors. Cardiopulmonary fitness and adherence results were reported(16); yet glucoregulatory dynamics remained unexplored.

Using *The Expecting Study* data, we evaluated intervention effects on glucoregulatory and inflammatory markers in this population. We hypothesized that the exercise intervention group (EX) would exhibit improved glucoregulatory and inflammatory profiles compared to the Standard-of-Care (SoC) control group, thereby mirroring the insulin-sensitizing effects of exercise observed in non-pregnant adults with obesity. Using continuous glucose monitoring (CGM) data from a participant subset, we explored how exercise affected maternal glucose variability and whether CGM metrics were associated with maternal inflammatory markers.

## Methods

### Data & Participants

This secondary study analyzed antepartum data from a RCT (https://clinicaltrials.gov/study/NCT02125149) conducted at the Arkansas Children’s Nutrition Center, Little Rock, AR, from September 2014 to May 2024. One participant with completed baseline procedures failed post-baseline visit eligibility review and was dropped. The analytic dataset included 263 eligible participants from the parent RCT (**Figure 1**), with n = 214 contributing to the final analysis. Study procedures and power estimation for the parent trial have been described(16). There was no public or patient involvement in the design, conduct, and reporting of this study.

**Figure 1.**
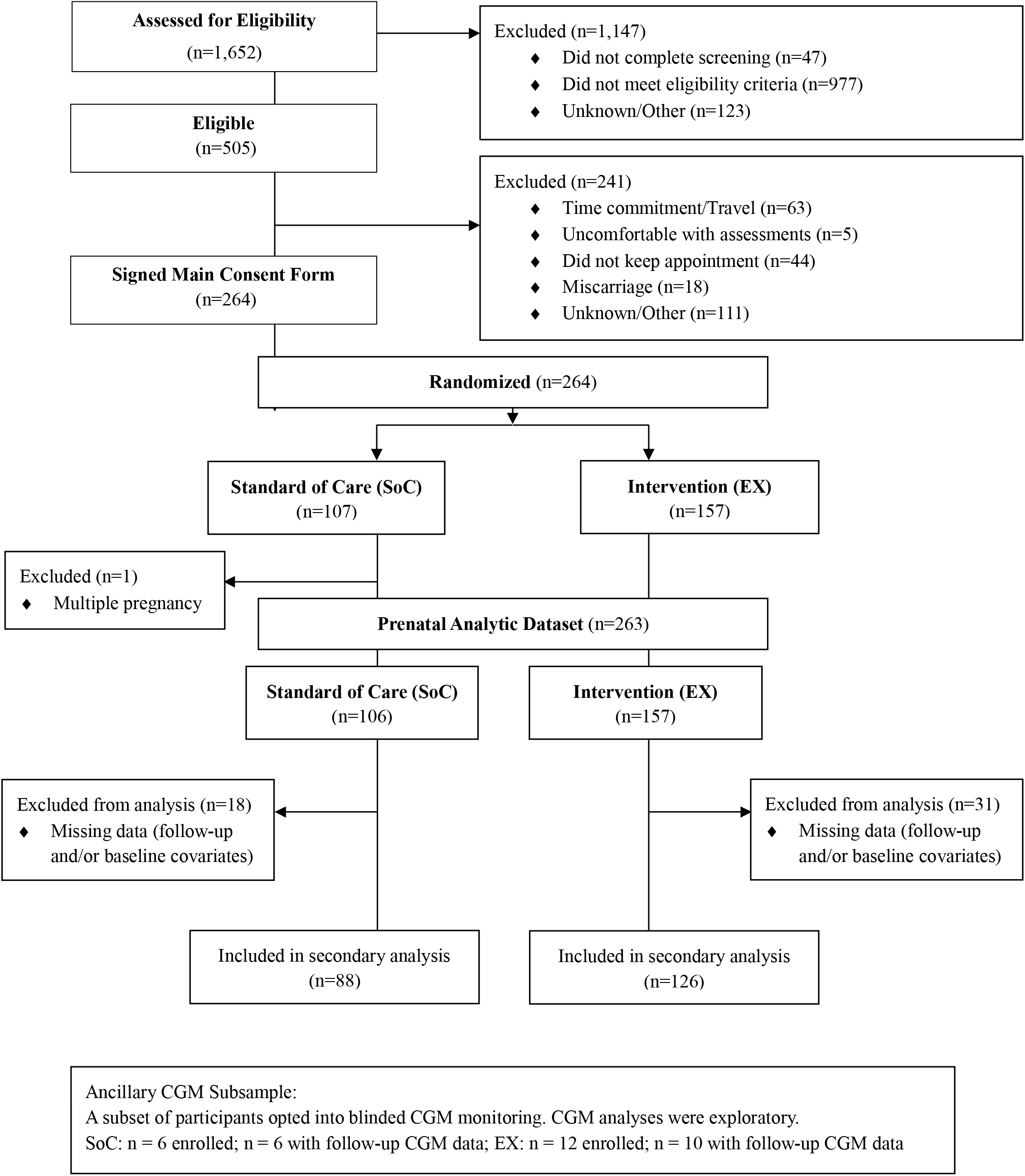
Consort Flow Diagram

In brief, women were enrolled by 12 weeks’ gestation. Eligibility criteria assessed via phone screening and confirmed during the baseline visit included: age ≥18 years, self-reported pre-pregnancy BMI ≥30.0 kg/m^2^, singleton pregnancy conceived without fertility treatments, and reporting a sedentary lifestyle (i.e., no structured physical activity engagement and sedentary occupational activity level). Exclusion criteria included multiple pregnancy, preexisting cardiometabolic conditions (e.g., diabetes or chronic hypertension), substance use, or contraindications to exercise during pregnancy.

Following enrollment and the baseline visit, participants were randomly assigned to either EX or SoC. Starting at gestational week 14.0 ± 0.8, EX completed progressive aerobic, resistance, and stretching sessions three times per week, designed to improve participants’ fitness levels in line with guidelines from the American College of Obstetricians and Gynecologists and the American College of Sports Medicine for exercise during pregnancy with obesity(17, 18). Outcome assessments took place once per trimester, at gestational weeks 12.14 ± 0.50, 24.19 ± 0.68, and 36.03 ± 0.58. Study procedures are described in detail in Holmes et al(16). Outcome assessment research team members and data analysts were blinded to intervention assignment. Scheduling and physical activity research core members were unblinded to facilitate the intervention protocol during center visits, separate from assessment visits. A protocol schematic is shown in **Supplemental Figure 1**.

All participants provided written informed consent for participation and received obstetric care provider clearance before enrollment. The protocol was approved by the University of Arkansas for Medical Sciences Institutional Review Board (#202954) in accordance with the Declaration of Helsinki.

### Blood Collection for the Quantification of Glucoregulatory and Inflammatory Markers

Following an overnight fast, antecubital venous blood samples were collected into serum and EDTA plasma vacutainers (Becton, Dickinson & Company, Franklin Lakes, NJ). Insulin, C-peptide, glucagon, interleukin-6 (IL-6), C-reactive protein (CRP), and tumor necrosis factor alpha (TNF-α) were quantified using an electrochemiluminescence detection platform, run on the MESI QuickPlex SQ 120 Imager (Meso Scale Diagnostics LLC, Rockville, Maryland) using commercially available kits (V-PLEX Metabolic Panel 1, V-PLEX Proinflammatory Panel 1 (human), V-PLEX Vascular Injury Panel 2 (human)). Serum glucose concentrations were determined using clinical analyzers (RX Daytona, Randox Laboratories Limited, Kearneysville, WV, USA; Roche COBAS Integra 400 Plus, Roche Diagnostics Corp, Indianapolis, IN, USA).

### Insulin Resistance and Beta-Cell Function Estimation

Fasting glucose (mmol/L), insulin (pmol/L), and C-Peptide (nmol/L) concentrations were used to calculate HOMA2-derived indices of insulin resistance (HOMA2-IR), insulin sensitivity (%S), and β-cell function (%B) using the HOMA2-IR Calculator macro (v2.2.3, Diabetes Trial Unit, University of Oxford, United Kingdom). Insulin-derived values were used for %S, while C-peptide-derived indices were reported for HOMA2-IR and %B, given that C-peptide, not subject to hepatic extraction, provides a more stable measure of endogenous insulin secretion(19).

### CGM Application and Wear-time Procedures

From October 2022 onward, participants opted in to an ancillary study using blinded CGM once per trimester. This component evaluated the feasibility of repeated CGM use in pregnancies with obesity but without diabetes and captured longitudinal glucose patterns while minimizing reactivity. Due to the later initiation of the ancillary study and the ongoing accrual of the parent cohort, the participant subset included in this analysis was not selected using stratified methods and may not provide a balanced representation across key demographics. Therefore, all CGM-related analyses reported herein are exploratory. Participants opting into the ancillary study wore CGM sensors (Freestyle Libre Pro, Abbott Diabetes Care, Alameda, CA, USA) once per trimester, at gestational weeks 11.90 ± 0.49, 24.50 ± 0.90, and 35.97 ± 0.31, respectively. Sensors worn on the back of the upper arm recorded interstitial glucose every 15 minutes for up to 14 days (1344 total possible observations per participant), with participants blinded to CGM data to minimize reactivity. Sensor data was downloaded to a de-identified cloud-based platform (*LibreView*) and used solely for research purposes. All participants were advised to undergo gestational diabetes screening per standard of care.

### CGM Wear-Time Metrics

CGM wear-time metrics characterized sensor adherence and data completeness **3**.

Metrics were computed per participant within each trimester and summarized as group means (SD). *Days worn* represented the number of unique calendar days with any CGM data recorded (00:00 – 23:59h). *Valid days* were defined as days with available CGM data and no evidence of sensor dropout, e.g., missing glucose data or recorded glucose values of 40 mg/dL. Values at or below 40 mg/dL were considered implausible or below device-detectable limits and removed prior to glycemic metrics calculations. While only 0.7% (393 of 53,025 total observations) met this removal criterion, these values could represent either a technical artifact or true low glucose events. Their removal may result in slightly inflated glycemic metrics and underestimated time below range (TBR).

*Mean daily wear time* was calculated as the average hours of CGM data captured per day, derived from the total number of 15-minute sensor readings divided by days worn. *Percent time* observed was calculated as the percentage of expected CGM observations within the wear window that were valid. Missing CGM data were not imputed given CGM metrics were averaged across multiple days. Multiple imputation by chained equations sensitivity checks will be made available at https://osf.io/egkx7 within 3 months of publication.

### CGM Metrics

Antepartum glycemic patterns were calculated from common clinically referenced CGM metrics(20) using iglu (version 4.3.0)(21). Values for *24-h mean glucose* were derived from all valid CGM readings across the 24-hour period. *Daytime mean glucose* was calculated from readings between 06:00h and 22:00h, and *nighttime mean glucose* from readings between 22:01h and 05:59h. *Time in Range (TIR)* was defined two-fold, given the lack of consensus for a ‘healthy range’ in this population: as 63 – 140 mg/dL (TIR_63-140_) per the American Diabetes

Association Standards of Care(22), and 63-120 mg/dL (TIR_63-120_) as proposed by Carlson et al(23), when describing normoglycemic pregnancies. Each TIR outcome reported the total hours per day during which CGM glucose values fell within this range. In a similar format, *Time Below Range (TBR)* and *Time Above Range (TAR)* were derived, as total hours per day CGM glucose values recorded either <63 mg/dL (TBR), >140 mg/dL (TAR_140_), or >120 mg/dL (TAR_120_). *Mean Amplitude of Glycemic Excursion (MAGE)* was derived using the validated, open-source algorithm by Fernandes et al on default settings(24). To explore intraday glycemic variability during specific time periods, Continuous Overall Net Glycemic Action [CONGA(4)] was calculated using glucose values paired 4 hours apart, within each window of interest(25), averaged across wear time by participant and trimester. All cleaned CGM data were used to derive participant-level daily metrics, which were analyzed using mixed-effects models to estimate marginal means by group and trimester.

### Statistical Approach

Baseline demographic and clinical characteristics were summarized as mean ± SD for continuous variables, and frequencies (%) for categorical variables. Because the CGM subset, was a convenience sample of the parent cohort, baseline between-group differences were assessed using Welch’s two-sample *t*-tests and Fisher’s exact tests. Longitudinal maternal glucoregulatory and inflammatory indices were analyzed using linear mixed-effects models with participant-specific random intercepts to account for within-subject correlation across repeated measurements. Fixed effects included group (EX vs. SoC), time (24-week vs. 36-week), and their interaction with adjustment for baseline (12-week) values, maternal age, BMI, and cardiorespiratory fitness. Analyses included participants with follow-up outcome data and complete baseline covariates. Models were fitted separately for the main cohort and CGM subgroup. Main and interaction effects were evaluated using Type II Wald Χ^2^ tests. Except for glucose, outcomes were log-transformed to improve residual normality. Model estimates were back-transformed and reported as symmetric percent changes (sympercent; s%).

CGM analyses were conducted as a post-hoc exploratory aim in an available subset with complete CGM data at multiple timepoints. No a priori power analysis was conducted; therefore, coefficients were interpreted cautiously, with confidence interval width used to estimate precision. CGM metrics were analyzed using the same linear mixed-effects model structure described above, including adjustment for baseline CGM values. Model assumptions were visually evaluated using residual and Q-Q plots. Statistical significance was set at α = 0.05, with no false discovery rate (FDR) or multiple comparison correction. Exact p-values (truncated at p < 0.001) are reported.

Linear mixed-effects models are robust to data missing at least at random. Analyses were performed in R (version 4.5.1)(26). Reporting followed the CONSORT guidelines(27).

## Results

### Participant Baseline Characteristics

Baseline demographic and anthropometric characteristics of participants are presented in **Table 1**. Per primary outcome design, participants were unequally randomized to EX, n = 157 or SoC, n = 106, with EX, n = 126 and SoC, n = 88, contributing complete baseline and follow-up data. Age, race/ethnicity, and key anthropometric measures were similar between groups. At 36 weeks, 17 participants reported a gestational diabetes diagnosis (GDM; EX, n = 7; SoC, n = 10).

**Table 1.** Participant Demographics.

|  | Main Cohort |  | CGM Subsample<sup>1</sup> |  |
| --- | --- | --- | --- | --- |
| Characteristic | SoC<br>N=88 | EX<br>N=126 | SoC<br>N=6 | EX<br>N=12 |
| Race |  |  |  |  |
| African American | 33 (37.5%) | 42 (33.3%) | 1 (16.7%) | 2 (16.7%) |
| Asian | 1 (1.1%) | 1 (0.8%) | -- | 1 (8.3%) |
| More than one Race | 1 (1.1%) | 1 (0.8%) | -- | -- |
| Unknown or Not Reported | 0 (0.0%) | 0 (0.0%) | -- | -- |
| White | 53 (60.2%) | 82 (65.1%) | 5 (83.3%) | 9 (75.0%) |
| Ethnicity |  |  |  |  |
| Hispanic | 4 (4.5%) | 5 (4.0%) | -- | -- |
| Non-Hispanic | 84 (95.5%) | 121 (96.0%) | 6 (100%) | 12 (100%) |
| Measurements at 12 weeks, mean (SD) |  |  |  |  |
|  | SoC<br>N=106 | EX<br>N=157 | SoC<br>N=6 | EX<br>N=12 |
| Age, years | 29.34 (4.86) | 30.07 (5.08) | 30.67 (2.76) | 31.38 (5.02) |
| Height, cm | 162.98 (6.24) | 164.43 (6.67) | 160.53 (5.0) | 161.14 (5.39) |
| Weight, kg | 100.69 (20.63) | 99.71 (15.48) | **83.64 (10.98)** | **95.80 (9.68)** |
| BMI, kg/m<sup>2</sup> | 37.81 (6.81) | 36.83 (4.92) | **32.38 (3.05)** | **36.97 (4.12)** |
| Body Fat Ratio | 0.48 (0.06) | 0.48 (0.05) | **0.43 (0.05)** | **0.49 (0.05)** |
| VO<sub>2</sub> (ml·kg<sup>-1</sup>·min<sup>-1</sup>) at RPE-15 | 16.54 (3.15) | 16.20 (2.86) | 17.39 (4.03) | 16.30 (3.03) |
| Data are n (%) unless otherwise indicated. 263 participants were randomized; a subsample of 18 participants opted into the CGM ancillary study.<br><sup>1</sup>Between-group differences at baseline were assessed via Welch's two-sample t-tests (continuous variables) and Fisher's exact test for categorical variables for the CGM subsample only because it was a convenience sample of the main cohort. **Bold font** denotes statistically significant group difference at baseline.<br>SoC: Standard of Care group; EX: Exercise group; RPE-15: Borg's Rating of Perceived Exertion of 15 measured during a submaximal fitness test; CGM: Continuous Glucose Monitoring; BMI: Body Mass Index; VO<sub>2</sub>: Oxygen consumption |  |  |  |  |

Eighteen participants (EX, n = 12; SoC, n = 6) opted into the ancillary CGM study. Because enrollment was opportunistic, the EX group exhibited higher baseline weight (mean difference,12.16 kg, 95% CI: [0.22, 24.1]), BMI (4.59 kg/m^2^, 95% CI: [0.88, 8.3], and body fat ratio (0.05, 95% CI: [0.00, 0.11]) compared to SoC (all p < 0.05). Race/ethnicity distributions were comparable between groups in the main cohort and CGM subgroup. Two CGM participants, both SoC, reported a GDM diagnosis at 36 weeks.

### Glucoregulatory Serum Marker Trajectories Across Pregnancy (Main Cohort)

#### Fasting glucose

No group, trimester, or group-by-trimester effects were observed for fasting glucose (all p ≥ 0.681), indicating stable glucose concentrations from 24 to 36 weeks across groups.

#### Other glucoregulatory markers

No group effects or group-by-trimester interactions were observed for the remaining glucoregulatory markers (all p > 0.132; **Table 2**). Significant trimester effects were detected for all remaining glucoregulatory outcomes (p<0.001), except glucagon (p = 0.096). From 24 to 36 weeks, *fasting insulin* concentrations rose by 16.3% (95% CI: [9.2%, 23.4%]; p < 0.001), *C-peptide* concentrations rose by 20.2% (95% CI: [11.4%, 28.9%]; p < 0.001), *HOMA2-IR* increased by 19.8% (95% CI: [11.1%, 28.6%]; p < 0.001), and *HOMA2-%B* (estimated β-cell function) increased by 15.2% (95% CI: [8.4%, 22.0%]; p < 0.001), whereas *HOMA2-%S* (estimated insulin sensitivity) decreased by 18.0% (95% CI: [-23.9%, - 12.2%]; p < 0.001). Estimated insulin sensitivity was approximately half, whereas estimated β-cell function was approximately twice the reference value, indicating pregnancy-associated insulin resistance. Glucagon concentrations showed no trimester or group-by-trimester effect (p = 0.096 and p = 0.132, respectively).

**Table 2.** Model-Estimated Marginal Means by Group and Trimester and Overall Effects from Adjusted Linear Mixed-Models ^3^.

| Model-Estimated Marginal Means (95% CI)<sup>1</sup> |  |  |  |  | Average Overall Treatment & Time Effects<sup>2</sup> |  |  |  |
| --- | --- | --- | --- | --- | --- | --- | --- | --- |
| Variable | 24 Weeks |  | 36 Weeks |  | Overall Treatment Effect<sup>3</sup> |  | Overall Time Effect<sup>3</sup> |  |
|  | SoC | Ex | SoC | Ex | Estimate<br>[95% CI] | p-value | Estimate<br>[95% CI] | p-value |
| **N = 214** | N = 88 | N = 126 | N = 88 | N = 126 |  |  |  |  |
| Glucose,<br>mg/dL | 88.02<br>[85.89, 90.15] | 88.53<br>[86.78, 90.28] | 87.81<br>[85.68, 89.93] | 88.25<br>[86.41, 90.10] | 0.478<br>[-1.802-2.757] | 0.681 | -0.245<br>[-1.863-1.372] | 0.766 |
| **N = 214** | N = 88 | N = 126 | N = 88 | N = 126 |  |  |  |  |
| Insulin,<br>(pmol/L) | 91.26<br>[83.08, 100.25] | 98.31<br>[90.99, 106.22] | 108.13<br>[98.47, 118.75] | 114.92<br>[105.93, 124.67] | 0.068<br>[-0.033-0.168] | 0.188 | **0.163**<br>**[0.092-0.234]** | **<0.001** |
| **N = 213** | N = 87 | N = 126 | N = 87 | N = 126 |  |  |  |  |
| C-Peptide,<br>(ng/mL) | 3.70<br>[3.35, 4.09] | 3.53<br>[3.25, 3.83] | 4.30<br>[3.89, 4.75] | 4.54<br>[4.17, 4.95] | 0.003<br>[-0.095-0.100] | 0.956 | **0.202**<br>**[0.114-0.289]** | **<0.001** |
| **N = 213** | N = 87 | N = 126 | N = 87 | N = 126 |  |  |  |  |
| Glucagon,<br>(pg/mL) | 4.00<br>[3.05, 5.25] | 5.20<br>[4.15, 6.50] | 5.22<br>[3.98, 6.85] | 5.35<br>[4.25, 6.74] | 0.143<br>[-0.177-0.462] | 0.382 | 0.148<br>[-0.007-0.303] | 0.061 |
| **N = 214** | N = 88 | N = 126 | N = 88 | N = 126 |  |  |  |  |
| HOMA2-IR<br>(C-Peptide) | 2.67<br>[2.41, 2.95] | 2.54<br>[2.34, 2.76] | 3.09<br>[2.80, 3.42] | 3.25<br>[2.98, 3.55] | 0.487<br>[-0.098-0.100] | 0.980 | **0.198**<br>**[0.111-0.286]** | **<0.001** |
| **N = 214** | N = 88 | N = 126 | N = 88 | N = 126 |  |  |  |  |
| HOMA2 %S<br>(Insulin) | 62.35<br>[57.25, 67.90] | 55.83<br>[52.06, 59.88] | 50.51<br>[46.40, 54.97] | 48.04<br>[44.64, 51.69] | -0.08<br>[-0.175-0.014] | 0.097 | **-0.180**<br>**[-0.239 - -0.122]** | **<0.001** |
| **N = 214** | N = 88 | N = 126 | N = 88 | N = 126 |  |  |  |  |
| HOMA2 %β<br>(C-Peptide) | 199.94<br>[185.01, 216.08] | 193.61<br>[181.65, 206.35] | 223.39<br>[206.76, 241.35] | 234.76<br>[219.50, 251.08] | 0.009<br>[-0.067-0.084] | 0.821 | **0.152**<br>**[0.084-0.220]** | **<0.001** |

<sup>1</sup> Values in the 24- and 36-week columns represent model-estimated marginal means (95% CI) from linear-mixed effects models with a random intercept for participant.
<sup>2</sup> Type II Wald X<sup>2</sup> tests were used to evaluate trimester, group, and group-by-trimester effects. No significant interactions were detected for any glucoregulatory marker and were omitted for clarity.
<sup>3</sup> Overall average treatment effects (EX-SoC) and average overall time effects (36-24 weeks) estimates are presented as symmetric percent change (s%) with 95% confidence intervals for log-transformed outcomes. Glucose is presented as absolute differences.
<sup>4</sup> All models included a random intercept for participant and were adjusted for baseline (12 weeks), maternal age, BMI, and cardiorespiratory fitness.
**Bold font** denotes statistically significant group difference at baseline.

### CGM metrics

#### Wear-time metrics

CGM adherence was high, averaging 12.3 ± 3.5 – 15.0 ± 0.0 days per trimester, with 99.3% valid readings from available CGM data (**Table 3**).

**Table 3.** Continuous Glucose Monitoring (CGM) Wear Time Metrics.

| Characteristic | 12 Weeks |  | 24 Weeks |  | 36 Weeks |  |
| --- | --- | --- | --- | --- | --- | --- |
|  | SoC | EX | SoC | EX | SoC | EX |
|  | N=6<sup>1</sup> | N=9<sup>1</sup> | N=5<sup>1</sup> | N=9<sup>1</sup> | N=6<sup>1</sup> | N=8<sup>1</sup> |
| Days Worn<sup>2</sup> | 14.8<br>(0.4) | 14.0<br>(1.7) | 15.0<br>(0.0) | 14.9<br>(0.3) | 12.3<br>(3.5) | 12.6<br>(4.5) |
| Valid Days<sup>3</sup> | 12.7<br>(3.3) | 13.7<br>(1.9) | 13.8<br>(2.2) | 13.8<br>(1.3) | 11.0<br>(4.8) | 12.6<br>(4.5) |
| Mean Wear Time Data Available<sup>4</sup><br>(hours/day) | 22.3<br>(0.3) | 22.2<br>(0.5) | 22.3<br>(0.0) | 21.8<br>(0.8) | 22.1<br>(0.3) | 21.7<br>(1.2) |
| % Time Observed<sup>5</sup> | 96.5<br>(5.4) | 99.8<br>(0.4) | 99.4<br>(1.3) | 99.7<br>(0.4) | 99.5<br>(0.7) | 100.0<br>(0.0) |

<sup>1</sup>Metrics are computed per participant within each trimester and summarized as Mean (SD) across participants within group.
<sup>2</sup>Days Worn: Number of unique calendar days with any CGM data recorded (00:00 – 23:59).
<sup>3</sup>Valid Days: Days worn excluding days flagged as invalid due to sensor dropout (any reading of $\leq 40$ mg/dL on that day).
<sup>4</sup>Mean Wear Time Data Available: Average hours of CGM data captured per day; calculated from the total number of records (15-min intervals) divided by days worn.
<sup>5</sup>Percent Time Observed: Proportion of CGM data that are observed (i.e., not missing/invalid) within the wear time window, expressed as percentage.

#### 24-hour mean glucose

The average 24-h mean glucose remained <100 mg/dL across groups and trimester (**Table 4**). A group-by-trimester interaction was observed (Χ*^2^* = 5.29, p = 0.021), average 24-hour mean glucose decreased from 24 to 36 weeks in SoC (Δ = −7.05 mg/dL; 95% CI: [-12.35, −1.75]; p = 0.009) but remained stable in EX (Δ =1.71 mg/dL; 95% CI: [−3.58, 7.00]; p = 0.526). At 36 weeks, estimated 24-hour mean glucose was 82.48 mg/dL (95% CI: [76.24, 88.72]) for SoC and 89.71 mg/dL (95% CI: [82.93, 96.49]) for EX. When stratified by day- and nighttime periods, the interaction was significant only for nighttime glucose (Χ*^2^* = 6.41, p = 0.011). Nighttime glucose decreased in SoC (Δ = −8.23 mg/dL; 95% CI: [-14.43, −2.08], p = 0.009), but remained stable in EX ( Δ = 2.99 mg/dL; 95% CI: [-3.19, 9.16], p = 0.343).

#### Time in Range (TIR)

TIR within the pregnancy target of 63 – 140 mg/dL(28), showed a group-by-trimester interaction (Χ*^2^* = 9.96, p = 0.002). From 24 to 36 weeks, TIR_63-140_ declined in SoC ( Δ = −2.27 h/day; 95% CI: [-3.49, −1.06]; p < 0.001), but remained stable in EX (Δ = 0.49 h/day (95% CI: [-0.72, 1.71]; p = 0.426). This pattern held when applying TIR_63-120_ (interaction Χ*^2^* = 7.05, p = 0.008), which decreased in SoC (Δ = −2.24 h/day, 95% CI: [-3.55, −0.93]; p = 0.001) but not EX (Δ = 0.27 h/day, 95% CI: [-1.04, 1.58], p = 0.684; **Figure 2**). This reduction in TIR corresponded to increased *Time Below Range* (*TBR_<63_*) in SoC (Δ = 2.23 h/day; 95% CI: [0.89, 3.57], p=0.001) but not EX ( Δ = −0.58 hours; 95% CI: [-1.91, 0.76], p = 0.399; interaction Χ*^2^* = 8.461, p = 0.004). No group-by-trimester interactions were observed for *TAR_>140_* or *TAR_>120_*; participants spent < 2h/day above 120 mg/dL, and < 0.5h/day above 140 mg/dL.

**Figure 2.**
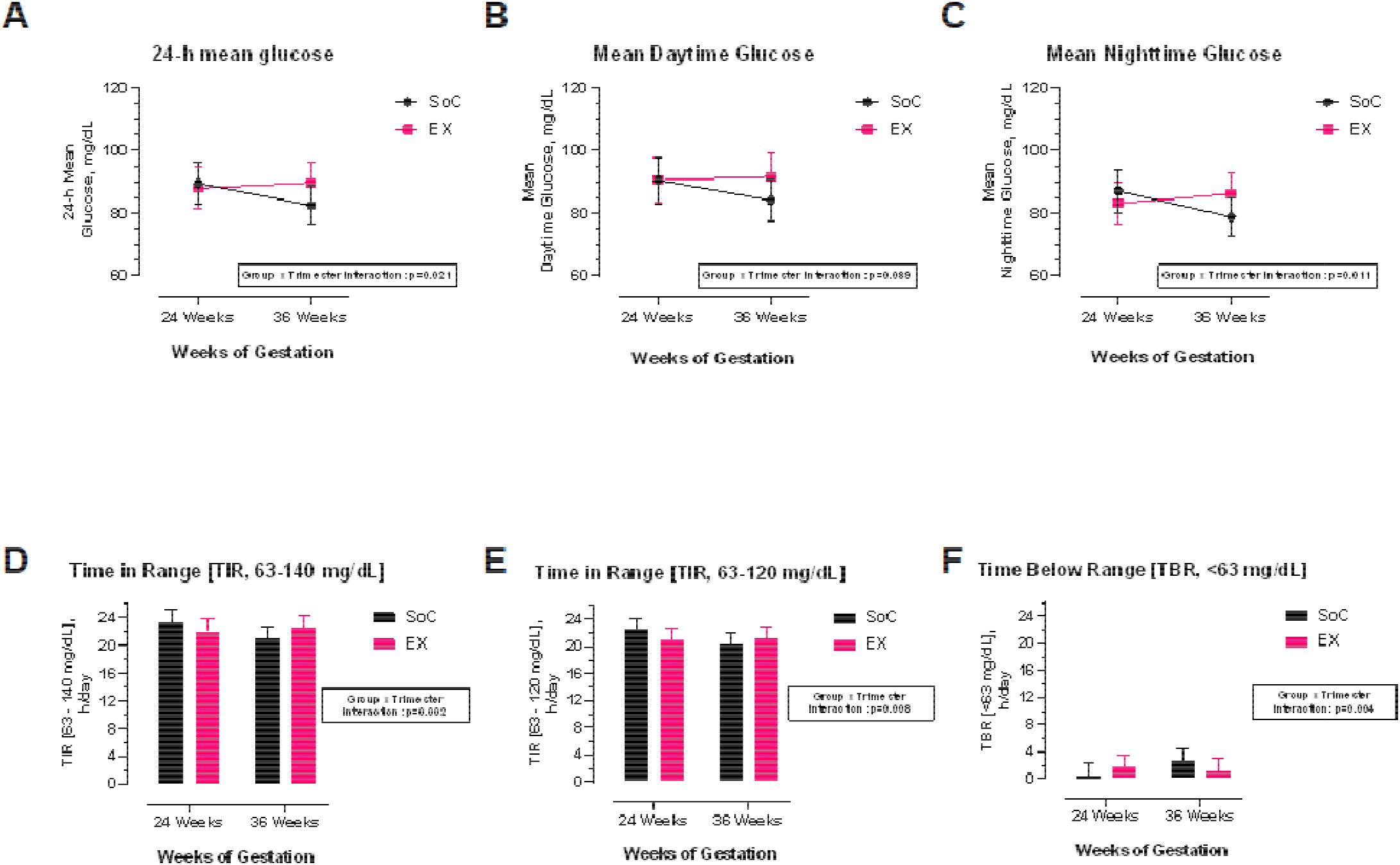
Continuous glucose monitoring summary metrics across pregnancy by study group. Model-estimated means ± SE for CGM-derived outcomes at 24 and 36 weeks’ gestation in women with obesity assigned to standard-of-care (SoC) or exercise (EX). Panels show (A) 24-h mean glucose, (B) mean daytime glucose, (C) mean nighttime glucose, (D) time in range (TIR, hours/day within 63-140 mg/dL), (E) TIR (hours/day within 63-120 mg/dL), and (F) time below range (TBR, hours/day <63 mg/dL). Group x Trimester interactions were significant for 24-h mean glucose (p = 0.021), mean nighttime glucose (p = 0.011), TIR 63-140 mg/dL (p = 0.002), TIR 63-120 mg/dL (p = 0.008), and TBR (p = 0.004). Models were adjusted for baseline values at 12 weeks, maternal age, BMI, and cardiorespiratory fitness.

#### Mean Amplitude of Glycemic Excursions (MAGE)

MAGE values did not differ between groups at 24 weeks (SoC, 37.10 mg/dL;EX, 41.67 mg/dL; Δ = 4.57 mg/dL (95% CI: [-2.19, 11.34), p = 0.185) or 36 weeks (SoC, 42.05 mg/dL; EX 42.12 mg/dL; Δ = 0.07 mg/dL (95% CI: [−6.35, 6.48], p = 0.984). No group, trimester, or group-by-trimester effects were observed (p ≥ 0.200), indicating stable MAGE across gestation in both groups.

#### Continuous Overall Net Glycemic Action [CONGA(4)]

During 16:00 – 20:00h (4 – 8pm), no group-by-trimester interaction was observed (p = 0.121), although trimester (Χ*^2^* = 6.59, p = 0.010) and group effects (Χ*^2^* = 3.83, p = 0.050) were detected. CONGA(4) increased from 24 to 36 in SoC (Δ = 3.07 mg/dL, 95% CI: [1.05, 5.09], p = 0.003) but did not change in EX. At 36 weeks, CONGA(4) was 3.51 mg/dL lower in EX vs SoC (95% CI: [-6.37, −0.65], p = 0.016).

Because the interaction was not significant, these contrasts do not provide robust evidence of differential change between groups. No significant group, trimester, or group-by-trimester effects were observed during 00:00-4:00 (all p > 0.170; **Table 4**).

### Maternal Inflammatory Markers

No group or group-by-trimester effects were observed for maternal IL-6, TNF-α, and CRP concentrations. All markers showed trimester effects (IL-6, Χ^2^ = 34.1; TNF-α, Χ^2^ = 16.65; CRP, Χ^2^ = 65.31; all p < 0.001). From 24 to 36 weeks, IL-6 concentrations increased by 17.6% (95% CI:[11.4, 23.8]) and TNF-α by 10.3% (95% CI: [5.2,15.5]), whereas CRP concentrations changed by −33.3% (95% CI: [-41.3, −25.3]; all p < 0.001; **Figure 3**).

**Figure 3.**
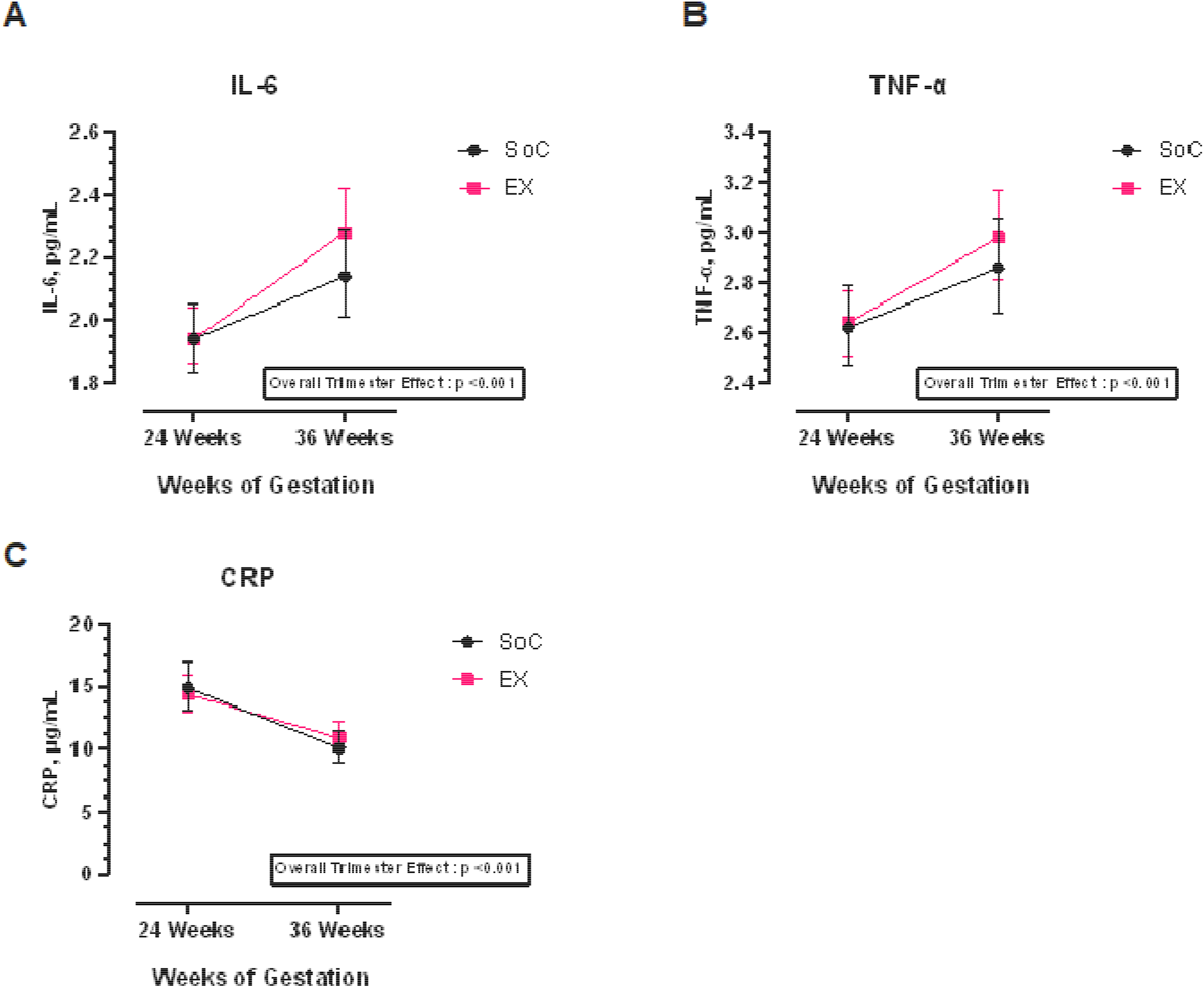
Maternal inflammatory markers across pregnancy by study group. Model-estimated means ± SE for maternal inflammatory markers at 24 and 36 weeks’ gestation in women with obesity assigned to standard-of-care (SoC, n=88) or exercise (EX, n=126). Panels show (A) IL-6, (B) TNF-α, and (C) CRP. There was a significant main effect for Trimester (all p<0.001). Models adjusted for baseline values at 12 weeks, maternal age, BMI, and cardiorespiratory fitness.

Cross-sectional associations of MAGE and TIR_63-140_ with inflammatory markers were examined separately at 24 and 36 weeks. At 24 weeks, greater TIR was associated with lower IL-6 (β = −0.23; 95% CI: [-0.43, −0.02]; p = 0.035), whereas neither CGM metric was associated with TNF-α or CRP. At 36 weeks, MAGE was positively associated with TNF-α (β=0.02; 95% CI: [0.00, 0.04]; p = 0.033). No other associations were observed. Group, BMI, age, and cardiorespiratory fitness were not associated with inflammatory markers at either time point (**Supplementary Table 1**).

## Discussion

This study examined whether initiating a structured, supervised, moderate-intensity exercise program altered fasting glucoregulatory and inflammatory trajectories across pregnancy in women with obesity, while also assessing the feasibility of blinded CGM-wear and monitoring of longitudinal glycemic patterns. In the main cohort, fasting glucose remained stable, whereas insulin, C-peptide, and HOMA2-indices showed the expected progression of insulin resistance and β-cell compensation from mid to late gestation, with no intervention effect. In the CGM subset, each monitoring period provided ≥ 12 days of data. CGM metrics generally remained within normoglycemic ranges in both groups. From 24 to 36 weeks, SoC exhibited modest declines in 24-h, nocturnal glucose and TIR, with increased TBR, whereas EX remained stable. Across monitoring periods, both groups spent < 20 min/day (1.4%) above 140 mg/dL, and approximately 100 min/day (6.9%) above 120 mg/dL. Neither MAGE nor CONGA(4) showed a significant group-by-trimester interaction, although afternoon CONGA(4) increased in SoC by 36 weeks. IL-6 and TNF-α increased, whereas CRP decreased across gestation. Associations of TIR or MAGE with IL-6 or TNF-α were small and trimester-specific.

This study extends the literature in three ways. First, it provides preliminary longitudinal evidence on intervention-related differences in 24-h mean glucose, time-in-range patterns, and inter- and intraday glycemic variability among pregnant women with obesity. Second, it examines exploratory within-trimester associations between CGM-derived glycemic variability and inflammatory markers. Third, it demonstrates the feasibility of integrating CGM into exercise-based pregnancy interventions outside diabetes monitoring.

The absence of group differences in fasting glucose, insulin, C-peptide, and HOMA2-indices suggests that late-gestation metabolic adaptations may outweigh modest effects of exercise initiated by 14 weeks’ gestation. This aligns with prior trials in women with overweight or obesity showing improved cardiorespiratory fitness and gestational weight gain without substantial changes in fasting glucose or insulin(29, 30). To our knowledge, this is the first large longitudinal study (N=214) to characterize HOMA2-estimated insulin sensitivity and β-cell function across gestation. Beta-cell function was approximately 1.9 to 2.3 times the reference level, whereas insulin sensitivity declined from approximately 0.6 to 0.5 times reference, consistent with progressive gestational insulin resistance. Glucagon findings warrant cautious interpretation. While the trimester effect and group-by-trimester interaction were not significant, within-group contrasts suggested increased glucagon in SoC but not EX. These findings are hypothesis-generating and do not establish that exercise attenuates gestational glucagon changes. The pattern is nevertheless biologically plausible given exercise-related modulation of hepatic glucose output and counterregulatory hormone(31). Larger RCTs with more frequent pre- and postprandial sampling are needed to clarify effects on glucagon-insulin dynamics in pregnancies with obesity.

While CGM has been proposed to support screening and prevention efforts in pregnancies at elevated risk for dysglycemia and adverse outcomes, its feasibility across at-risk populations and optimal target ranges remain uncertain(23, 32). Our study extends the literature by demonstrating high-adherence and feasibility data for blinded CGM during an antepartum exercise in pregnant women with obesity and by providing preliminary evidence that exercise may help preserve late-gestation glucose patterns and intraday variability.

In contrast to the null fasting findings, CGM captured subtler glycemic adaptations during gestation. Mean 24-h glucose and MAGE remained within normoglycemic ranges(33, 34), while TIR_63-140_ and TIR_63-120_ were comparable to values reported in uncomplicated pregnancies(23), consistent with preserved overall glycemic control despite obesity. However, significant group-by-trimester interactions indicated modest declines for 24-h, nighttime glucose, and TIR in SoC, whereas these metrics remained stable in EX. The nocturnal decline in SoC may reflect the increasingly pronounced overnight glucose nadir described in uncomplicated pregnancy(35).

Although differences assessed were modest, approximately 7 mg/dL for nocturnal glucose and 2h/day for TIR, sustained shifts in overnight glucose exposure may be clinically relevant in populations at elevated obstetric risk(36). While afternoon CONGA(4) also increased within SoC in late gestation, lack of significant interaction and concurrent insulin measurements limit mechanistic interpretation. Changes in meal timing, dietary intake, or activity patterns may have contributed, as even modest postprandial activity can alter 24-h glucose exposure in gestational diabetes(12). Although mean energy intake or physical activity did not differ between groups(16), time-of-day behaviors were not assessed. Larger, appropriately powered studies with greater separation in cardiorespiratory fitness and physical activity will be needed to confirm these patterns. Therefore, findings should be interpreted as exploratory signals that CGM can detect glycemic changes not captured by fasting markers, rather than robust evidence of exercise effects on afternoon or overnight glycemia in pregnancy.

The inflammatory profile observed was broadly consistent with pregnancy-related changes reported in women with obesity(37). Consistent with the trimester-specific immune and metabolic adaptations, IL-6 and TNF-α increased from 24 to 36 weeks(38). In contrast, CRP decreased, consistent with longitudinal findings in normal-weight pregnancies, but contrary to trends in CRP reported in pregnancies with obesity(37, 39). Given the dynamic, pleiotropic nature of these markers during pregnancy, attributing these changes to specific metabolic functions would be speculative(40).

Within-trimester associations between CGM metrics and inflammation were modest. At 24 weeks, greater TIR_63-140_ was associated with lower IL-6, suggesting that more time within the ADA-defined pregnancy target range may correspond to a less proinflammatory milieu in women with obesity, but without diabetes(28). At 36 weeks, greater MAGE was related to higher TNF-α, consistent with findings in non-pregnant women with obesity(7). Because effect sizes were small and associations were inconsistent across markers and timepoints, these findings should be considered preliminary.

This study has several strengths including the randomized design, supervised mixed-modality exercise intervention, repeated trimester-specific assessments of glucoregulatory and inflammatory markers, and integration of fasting biomarkers, HOMA2 indices, and CGM. Together, these measures provide a comprehensive characterization of gestational metabolic adaptations in women with obesity but without overt diabetes, while high CGM adherence supports the feasibility of blinded CGM in antepartum lifestyle interventions. However, limitations include the modest increase in physical activity despite improved cardiovascular fitness(16), which may have limited metabolic effects. The ancillary CGM subset was small, without stratified randomization; reducing precision and increasing susceptibility to baseline imbalance and type II error. Finally, enrollment of generally healthy women with obesity limits generalizability to more clinically diverse populations.

## Conclusion

In this randomized trial of pregnant women with obesity, supervised mixed-modality exercise initiated by the 14^th^ week of pregnancy did not significantly alter fasting glucoregulatory or inflammatory trajectories. In the exploratory CGM subset, glucose remained largely within normoglycemic ranges during monitoring. From 24 to 36 weeks, only SoC showed modest declines in 24-h, nocturnal glucose and TIR, with increased TBR, whereas EX maintained glycemic metrics. Associations between CGM metrics and inflammatory markers were small and inconsistent across gestational timepoints.

These findings suggest that moderate-intensity exercise may not alter pregnancy-related insulin resistance or inflammatory adaptations in this population, but may help preserve glycemic stability. High CGM adherence supports CGM as a feasible tool for evaluating antepartum lifestyle interventions.

## Acknowledgements

The authors would like to thank the study participants for their volunteer time and dedication. The authors acknowledge Lilian Cheak of the Arkansas Children’s Research Institute, and the ACNC Maternal and Early Nutrition for Optimal Growth and Development (MEND) Core for data collection, the ACNC data manager, Lindsey Fullen, and Staff Scientist Clark Sims, for data support, and the ACNC Physical Activity and Exercise Metabolism (PAEM) Core for exercise testing of the participants. L.T.J. would like to acknowledge the support provided during preparation of this manuscript by the Center for Childhood Obesity Prevention’s Scholarly Writing Program, funded by the National Institute of General Medical Sciences of the National Institutes of Health under Award Number P20GM109096 (Arkansas Children’s Research Institute, PI: Børsheim). A.W.B. is supported in part by the Arkansas Children’s Research Institute and the Arkansas Biosciences Institute. The views expressed in this article represent those of the authors and do not necessarily reflect the views of any organization.

## Data Availability Statement

Deidentified, individual participant data that underlie the results reported in this article, will be made available to inquiring researchers submitting a methodologically sound research proposal. Proposals should be directed to. Study protocol, statistical analysis plan, and analytic code will be made available beginning 3 months following article publication at https://osf.io/egkx7.

## Clinical Trial Registration

The Expecting Study – Prenatal Activity Intervention, NCT02125149

- 04/29/2026 First Posted
- 09/02/2014 Study Start https://clinicaltrials.gov/study/NCT02125149?term=NCT02125149&viewType=Card&rank=1

## Author Contributions

E.B., and A.A., conceived and designed the study; E.B., A.A., B.B., A.M. and L.T.J. performed experiments for current exploratory analyses. S.C., A.R.C., and A.W.B. performed statistical analyses. L.T.J. wrote the manuscript, and E.B., A.A., A.B.W., A.R.C.,S.C., B.B. and A.M. made major contributions to manuscript revisions. All authors read and approved the final manuscript. L.T.J., E.B., and A.A. are the guarantors of this work and take responsibility for the integrity of the data and accuracy of the data analysis.

