## Supplemental Table 1 for "Glucoregulatory and Inflammatory Trajectories in Pregnancy with Obesity: Secondary Analysis of a Randomized Exercise Trial"

|  | **24 WK** | | | | | | **36 WK** | | | | | |
| --- | --- | --- | --- | --- | --- | --- | --- | --- | --- | --- | --- | --- |
| **Predictors** | **Coefficients (95% CI)** | | | | | | **Coefficients (95% CI)** | | | | | |
|  | **IL-6 (pg/mL)** | ***p*** | **TNF-α (pg/mL)** | ***p*** | **CRP**  **(μg/mL)** | ***p*** | **IL-6 (pg/mL)** | ***p*** | **TNF-α (pg/mL)** | ***p*** | **CRP**  **(μg/mL)** | ***p*** |
|  | n = 13 |  | n = 13 |  | n = 13 |  | n = 13 |  | n = 13 |  | n = 13 |  |
| **Intercept** | **6.96 (0.61, 13.31)** | **0.036** | 3.32  (-4.55, 11.18) | 0.342 | 2.26 (-15.70, 20.22) | 0.769 | 2.29 (-6.29, 10.88) | 0.538 | -0.48 (-3.37, 2.41) | 0.698 | -5.29 (-19.63, 9.05) | 0.401 |
| **Group [EX]** | 0.33 (-0.27, 0.92) | 0.224 | 0.30 (-0.44, 1.03) | 0.358 | 0.06 (-1.62, 1.74) | 0.934 | 0.50 (-0.57, 1.57) | 0.294 | 0.30 (-0.06, 0.66) | 0.089 | -0.24 (-2.02, 1.54) | 0.750 |
| **BMI,**  **kg/m^2^** | -0.01 (-0.08, 0.05) | 0.681 | -0.04 (-0.12, 0.04) | 0.303 | 0.08 (-0.11, 0.26) | 0.359 | 0.03 (-0.08, 0.14) | 0.540 | -0.02 (-0.06, 0.02) | 0.295 | 0.09 (-0.10, 0.28) | 0.275 |
| **MAGE,**  **mg/dL** | -0.01 (-0.04, 0.01) | 0.258 | -0.01 (-0.04, 0.03) | 0.661 | 0.03 (-0.05, 0.11) | 0.374 | 0.00 (-0.06, 0.06) | 0.956 | **0.02 (0.00, 0.04)** | **0.033** | 0.04 (-0.06, 0.15) | 0.339 |
| **TIR,**  **hours/day** | **-0.23 (-0.43, 0.02)** | **0.035** | -0.12 (-0.37, 0.14) | 0.304 | -0.18 (-0.76, 0.40) | 0.480 | -0.11 (-0.34, 0.12) | 0.290 | -0.05 (-0.13, 0.03) | 0.151 | -0.07 (-0.45, 0.32) | 0.683 |
| **VO_2_ ,**  **(ml/kg/min)** | -0.07 (-0.16, 0.02) | 0.116 | 0.00 (-0.11, 0.11) | 0.979 | -0.06 (-0.32, 0.19) | 0.560 | 0.01 (-0.15, 0.17) | 0.911 | 0.05 (-0.01, 0.10) | 0.086 | 0.07 (-0.20, 0.34) | 0.543 |
| **Age,**  **Years** | -0.01 (-0.06, 0.04) | 0.659 | 0.01 (-0.05, 0.08) | 0.623 | 0.05 (-0.10, 0.20) | 0.467 | -0.05 (-0.20, 0.10) | 0.431 | 0.00 (-0.05, 0.05) | 0.833 | 0.09 (-0.16, 0.34) | 0.409 |

**Supplementary Table 1.** Within-Trimester Associations Between Maternal Inflammatory Markers and CGM Metrics

**Supplementary Table 1.** Coefficients (95% CI) and p-values from linear models examining associations between maternal characteristics, CGM metrics, and IL-6, TNF-α, and CRP at 24 and 36 weeks’ gestation. Models include study group, maternal BMI, MAGE, time in range [TIR], cardiorespiratory fitness [VO_2_], and maternal age. Biomarkers are reported in raw units for clinical interpretability.
