## Supplemental Figure 1 for "Glucoregulatory and Inflammatory Trajectories in Pregnancy with Obesity: Secondary Analysis of a Randomized Exercise Trial"

**Supplemental Figure 1.** Protocol Schematic

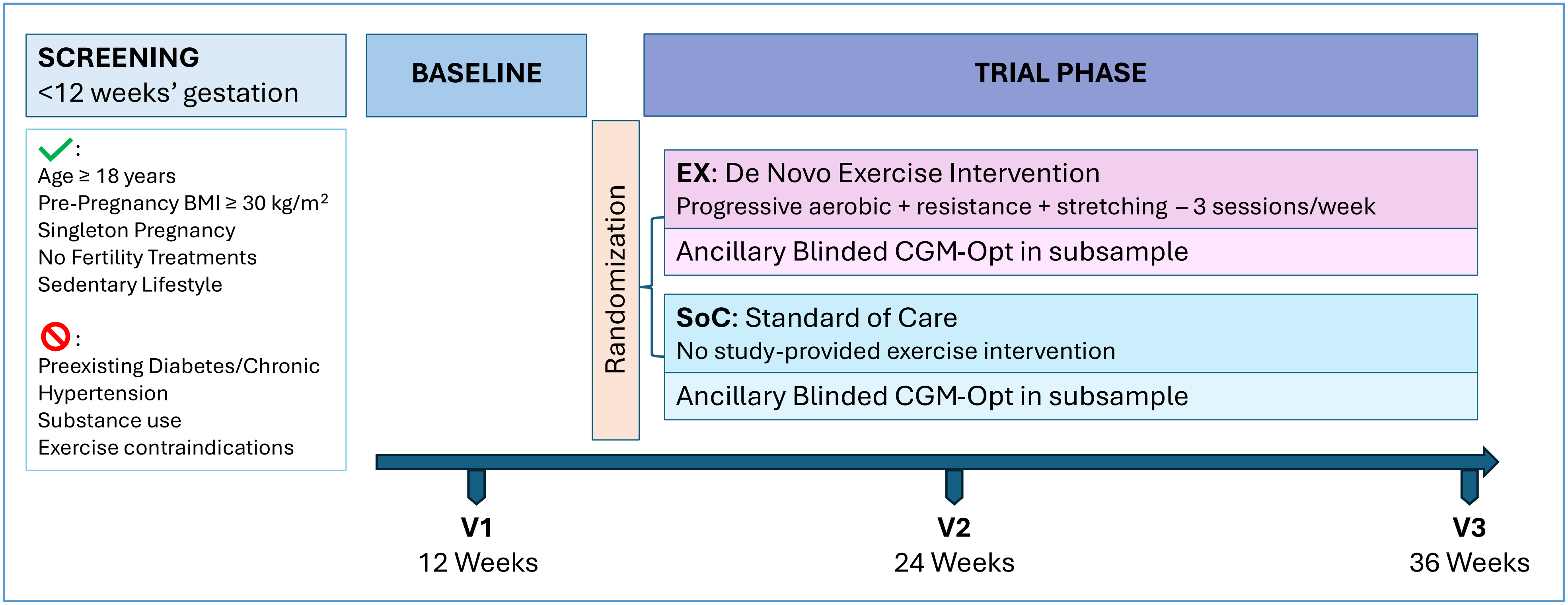


**Supplemental Figure 1.** Women with singleton pregnancies, pre-pregnancy BMI ≥ 30 kg/$m^2$, leading a sedentary lifestyle were enrolled at 12 weeks of gestation and randomized following the baseline visit to either an exercise intervention (EX) or standard-of-care control (SoC). The EX group began supervised progressive aerobic, resistance, and stretching exercise sessions 3x/week starting by 14 weeks' gestation. Outcome assessments were conducted once per trimester at approximately 12, 24, and 36 weeks' gestation. A subsample of participants opted into blinded continuous glucose monitoring (CGM) once per trimester as an exploratory ancillary study.
